# Comparing human vs. machine-assisted analysis to develop a new approach for Big Qualitative Data Analysis

**DOI:** 10.1101/2024.07.16.24310275

**Authors:** Sam Martin, Emma Beecham, Emira Kursumovic, Richard A. Armstrong, Tim M. Cook, Noémie Déom, Andrew D. Kane, Sophie Moniz, Jasmeet Soar, Cecilia Vindrola-Padros, collaborators

## Abstract

**Background:** Analysing large qualitative datasets can present significant challenges, including the time and resources required for manual analysis and the potential for missing nuanced insights. This paper aims to address these challenges by exploring the application of Big Qualitative (Big Qual) and artificial intelligence (AI) methods to efficiently analyse Big Qual data while retaining the depth and complexity of human understanding. The free-text responses from the Royal College of Anaesthetists’ 7th National Audit Project (NAP7) baseline survey on peri-operative cardiac arrest experiences serve as a case study to test and validate this approach.

**Methodology/Principal Findings:** Quantitative analysis segmented the data and identified keywords using AI methods. In-depth sentiment and thematic analysis combined natural language processing (NLP) and machine learning (ML) with human input - researchers assigned topic/theme labels and sentiments to responses, while discourse analysis explored sub-topics and thematic diversity. Human annotation refined the machine-generated sentiments, leading to an additional “ambiguous” category to capture nuanced, mixed responses. Comparative analysis was used to evaluate the concordance between human and machine-assisted sentiment labelling. While ML reduced analysis time significantly, human input was crucial for refining sentiment categories and capturing nuances.

**Conclusions/Significance:** The application of AI-assisted data analysis tools, combined with human expertise, offers a powerful approach to efficiently analyse large-scale qualitative datasets while preserving the nuance and complexity of the data. This study demonstrates the potential of this novel methodology to streamline the analysis process, reduce resource requirements, and generate meaningful insights from Big Qual data. The integration of NLP, ML, and human input allows for a more comprehensive understanding of the themes, sentiments, and experiences captured in free-text responses. This study underscores the importance of continued interdisciplinary collaboration among domain experts, data scientists, and AI specialists to optimise these methods, ensuring their reliability, validity, and ethical application in real-world contexts.

**Author Summary:** The use of Artificial intelligence (AI) in health research has grown over recent years. However, analysis of large qualitative datasets known as Big Qualitative Data, in public health using AI, is a relatively new area of research. Here, we use novel techniques of machine learning and natural language processing where computers learn how to handle and interpret human language, to analyse a large national survey. The Royal College of Anaesthetists’ 7th National Audit Project is a large UK-wide initiative examining peri- operative cardiac arrest. We use the free-text data from this survey to test and validate our novel methods and compare analysing the data by hand (human) vs. human-machine learning also known as ‘machine-assisted’ analysis. Using two AI tools to conduct the analysis we found that the machine-assisted analysis significantly reduced the time to analyse the dataset. Extra human input, however, was required to provide topic expertise and nuance to the analysis. The AI tools reduced the sentiment analysis to positive, negative or neutral, but the human input introduced a fourth ‘ambiguous’ category. The insights gained from this approach present ways that AI can help inform targeted interventions and quality improvement initiatives to enhance patient safety, in this case, in peri-operative cardiac arrest management.

## Introduction

The primary focus of this paper is to examine and compare human vs. human-machine learning (ML) enhanced analysis of a large qualitative dataset on the topic of peri-operative cardiac arrest. Through this comparison, we seek to assess the effectiveness and efficiency of machine-assisted analysis in relation to human analysis, while still capturing the valuable insights provided by human expertise in the context of patient safety research. We use the Royal College of Anaesthetists’ 7th National Audit Project (NAP7) survey to enable this comparison. The NAP7 methodology included two baseline surveys [1, 2], the second of which assessed knowledge, attitudes, practices and experiences of anaesthetists and anaesthesia associates relating to peri-operative cardiac arrest (S2 Appendix).

Big Qualitative (Big Qual) analysis of large free-text data using Artificial Intelligence (AI) techniques such as ML and natural language processing (NLP) shows an increased potential to handle large amounts of public health information [3]. Big Qual data is defined as datasets containing at least 100 participants, either as a stand-alone project or within a large quantitative study [4]. The ability to handle large amounts of data is particularly important in health research when there can be huge datasets from surveys or audit data and there is a need to rapidly analyse the results and make changes to clinical practice - e.g., during a pandemic when decisions need to be made quickly. The use of AI in health research has grown over recent years, recently culminating in the World Health Organisation (WHO) guidance document on the ethics and governance of AI for health [5]. However, the application of AI in Big Qual analysis public health datasets is still in its infancy, while the community works on defining appropriate methods, standards, regulations, and tools to use to analyse Big Qual data [6].

Patient safety and quality improvement in healthcare is one area that has seen benefits from the application of Big Qual research methods [7]. Researchers have employed these techniques to analyse data from sources such as social media and survey responses, identifying themes related to public health issues such as vaccine hesitancy in pregnant women [8]. Big Qual research methods, such as ML and NLP, are increasingly being applied to large healthcare datasets to gain insights and inform quality improvement efforts. While NLP offers the ability to rapidly analyse vast amounts of data, a criticism is that it may miss nuances captured by human qualitative analysis [9]. To address this, an approach combining NLP with expert human input, known as “machine-assisted topic analysis,” has been proposed [10]. While ML shows promise for classifying incident reports [11], we argue that a balance with manual processing remains crucial for a nuanced analysis.

The initial NAP7 survey with over 10,000 clinician responses provided a good data context to conduct a comparative analysis between human coding and AI methods [2]. In this paper we show how Big Qual analysis can support these quantitative findings by providing nuance and examples of anesthetists’ experiences. The study aims to apply machine-assisted sentiment, discourse, and thematic analysis to the free-text data from a large national survey of anesthetists (NAP7) on peri-operative cardiac arrest experiences [2] and compare against human-only analysis.

## Methodology

### Data preparation

The primary aim of this study was to compare the effectiveness and efficiency of machine-assisted analysis, using the AI tools Infranodus [12, 13] and Caplena [14], against human-only analysis in the context of qualitative data from the NAP7 Baseline Survey. By employing a comparative approach, we sought to assess the potential benefits and limitations of integrating AI tools into the qualitative research process, while maintaining the nuance and context provided by human researchers. This comparative methodology was applied throughout the data preparation, analysis, and interpretation stages of the study. The methodology for this study involved several steps (Fig 1). First, all 10,746 responses were downloaded from the SurveyMonkey® survey [15] to Excel. Two researchers, one digital analyst and one anaesthetist (SM1, EK), cleaned the data in Excel over two months. This process involved checking responses, creating a coding key to explain medical terms/slang for non-medical team members, adapting response categories, removing duplicates, and converting free-text responses for each question into a CSV file [2]. Researchers (EK and SM1) chose questions to analyse based on their relevance to the impact of peri-operative events on anaesthetists’ well-being and confidence. They chose six main questions (Q2, Q4, Q30, Q36, Q41 and Q43) that best answered the related themes and quantitative findings of the main paper [2]- giving a final sample of 5,196 responses. These included confidence levels in managing peri-operative cardiac arrest, insights into debriefing practices, having a rest break, or stopping work after being involved in a cardiac arrest, the prevalence of informal vs. formal well-being support and improvements needed in the post-cardiac arrest management or staff. Next, quantitative free-text analysis was performed using AI methods via the Pulsar Platform [16], where segmentation and keyword analysis were conducted to identify key topics of concern, key actors (e.g., segmentation by gender and stage in career), as well as word frequency analysis. These data were then used as a base template for the main thematic and sentiment analysis.

**Fig 1.**
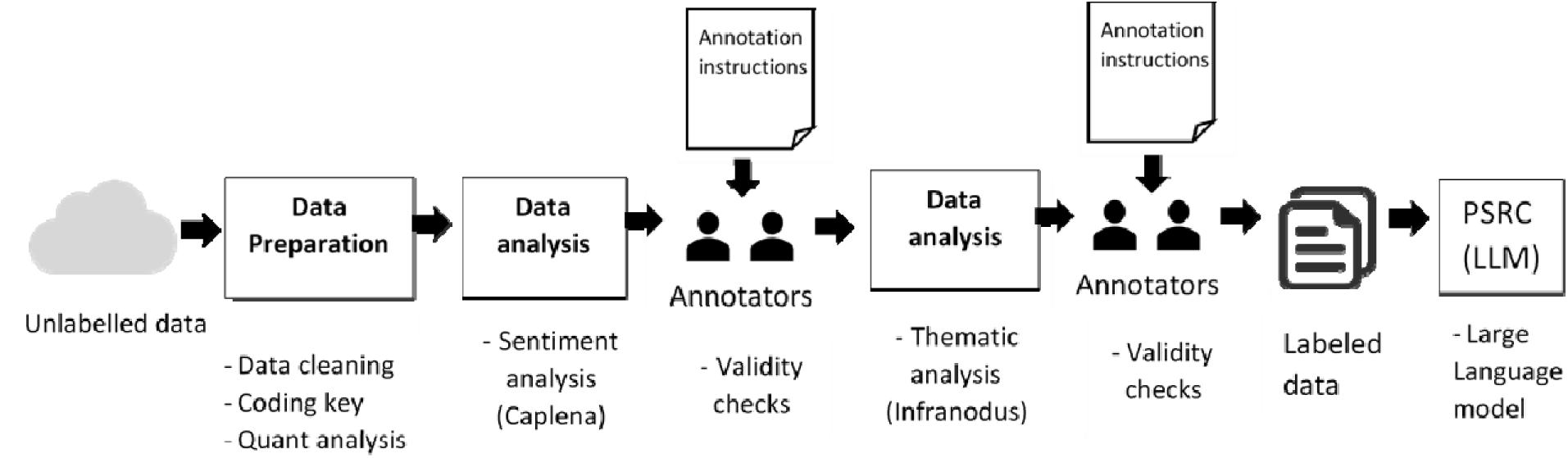
Human assisted machine learning approach. *PSRC = Patient Safety Research Collaboration, LLM = Large Language Model*.

We consulted the Standards for Reporting Qualitative Research (SRQR) guidelines to present the methodological process and the results [17].

### Data analysis-Natural Language Processing

Natural language processing (NLP) analytics [14] were combined with human input for sentiment analysis. Using Caplena, each response or statement was assigned a topic/theme label or sub- topic/theme. This meant that a response might have three sub-topics/themes attached to it. To train the Caplena model, five researchers including social scientists, a digital analyst, and an anaesthetist (EB, SM2, ND, SM1, EK,) reviewed a small subset of around 10% of the responses for each question, adding themes and sub-themes, revising labelling, and at times correcting labels. The model then learned from the changes made by the researchers and adjusted its labelling of the dataset to improve accuracy. Researchers documented the changes they made in each section before any subsequent changes to evaluate how well the model learned from human coding and to ensure consistency.

### Data analysis - Sentiment analysis

A sentiment analysis framework was established to categorise sentiments in the Baseline Survey responses (Fig 2). This framework guided the training of sentiment and thematic analysis tools. Three researchers (EB, SM1, EK) were involved in the initial manual annotation of the data, adding columns for the overarching sentiments: positive, negative, and neutral. The initial agreement among all three annotators was 80%. For the next stage, two researchers (EB and SM1) compared machine-generated sentiments with those initially assigned by the three human annotators. The agreement between these two main human annotators (EB, SM1) at this stage was 82%. After discussing and resolving discrepancies, the final agreement between EB and SM1 reached 100% for some themes and 92% for others. However, during the comparative analysis, with an in-depth reading of the responses - it became evident that the standard sentiment framework consisting of positive, negative, and neutral labels was insufficient to capture the nuances and complexities present in the free-text descriptions of anaesthetists’ experiences of cardiac incidents and their impact. The human annotators recognised the need for an additional sentiment label. This was initially labelled as “nuanced/mixed” [2], however, to ensure clarity and consistency in our analysis, we have revised the sentiment label to the name: “ambiguous” - as well as more clearly defined each sentiment in Fig 2 - to better represent the subtleties and ambiguities inherent in the text. Additionally, four columns (one by each of the four sentiments (positive, negative neutral and ambiguous) were added to the analysis spreadsheet for each researcher to assign a percentage to each sentiment option, allowing for a more granular representation of sentiment and acknowledging that a single response may contain elements of multiple sentiments in varying proportions. To aid in model building and ensure consistency, the researchers followed a specific protocol (Fig 2) when assigning sentiment percentages, requiring the assigned percentages to sum up to 100% for each response.

**Fig 2.**
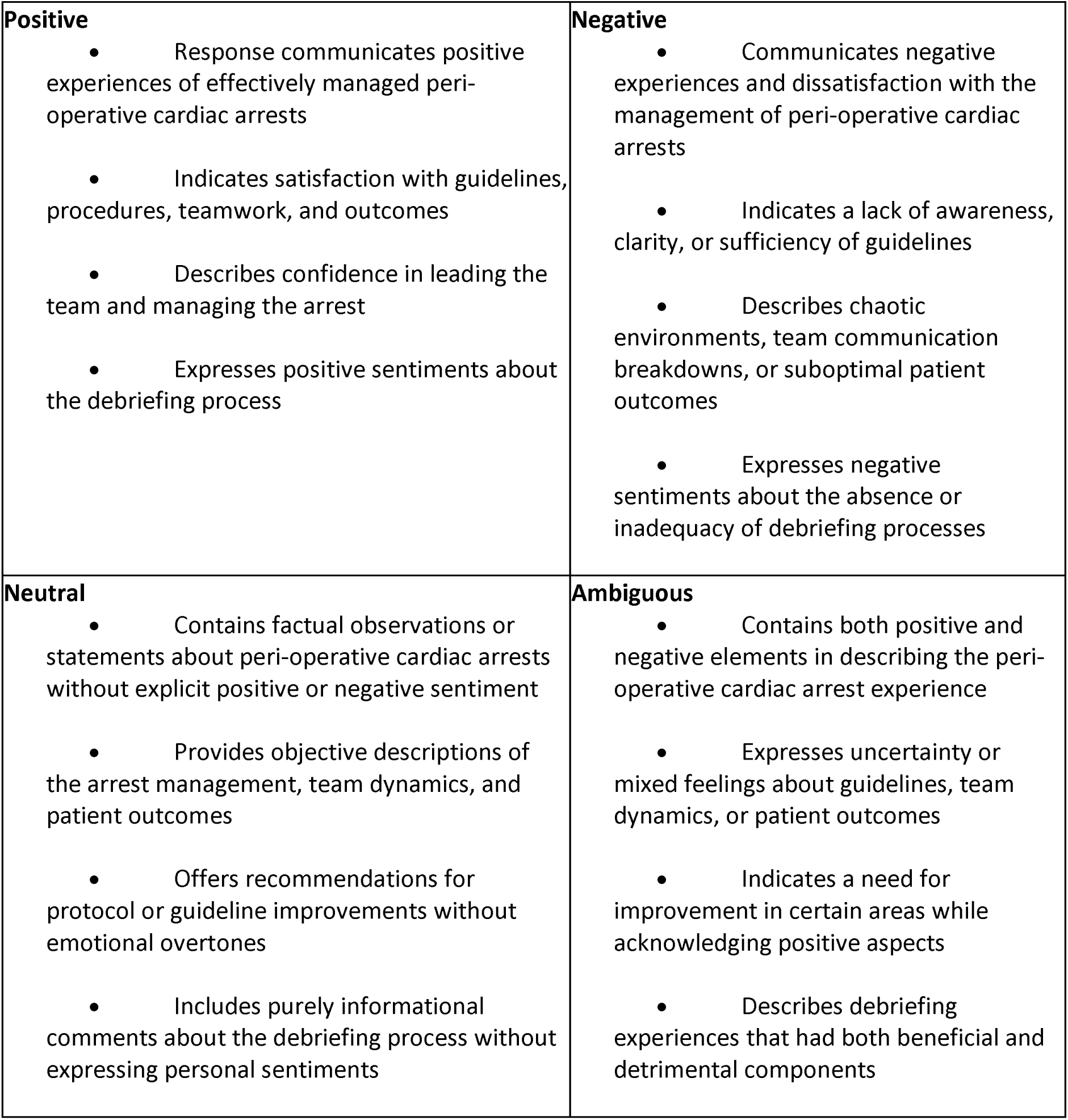
Sentiment Analysis Framework: Definitions of Positive, Negative, Neutral and Ambiguous sentiment – coded by human annotators.

### Data analysis - Sub-analysis: thematic and discourse analysis

A sub-analysis of the main themes identified with Caplena was conducted using the text network analysis tool Infranodus for discourse and thematic analysis, measuring themes and patterns around peri-operative cardiac arrest. The betweenness centrality of sub-topics was explored, which is a measure of how often a particular sub-topic acts as a bridge connecting different topic clusters. By analysing the connections between sub-topics, we were able to identify similarities in opinions and experiences expressed by participants, providing a more nuanced understanding of the relationships between various themes. Data was imported by question and theme into Infranodus to highlight keyword clusters of interest and the most influential topics. The distribution of topics across survey questions was measured where the most prominent relations between the nodes for each topic were represented by the closeness/betweenness of different clusters in relation to each other (Fig 3).

**Fig 3.**
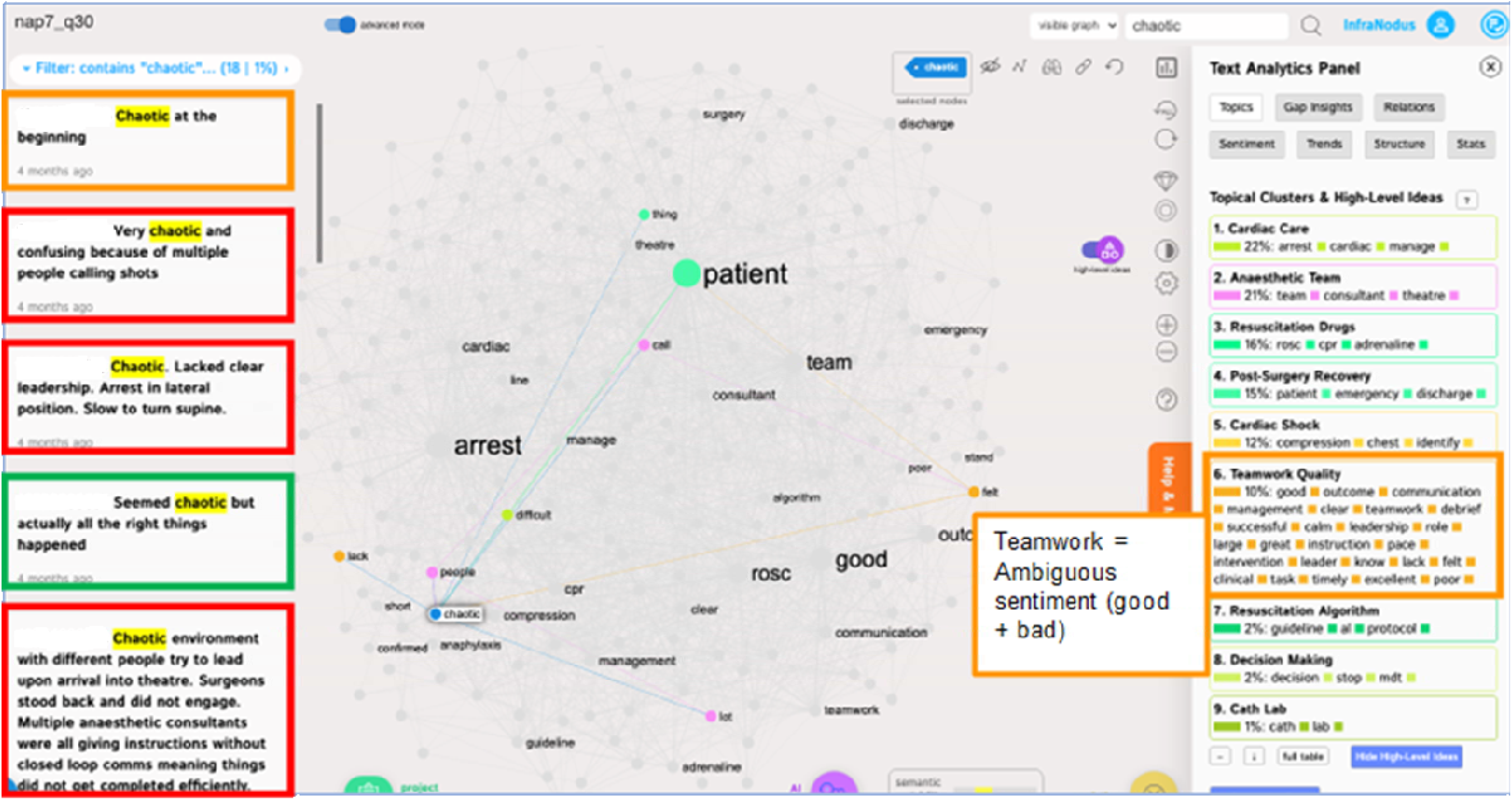
Infranodus: Example thematic network graph for the theme ‘chaotic’ within Q30-Satisfaction with quality of the management of the arrest. (Green boxes represent the positive sentiment statements, Red-negative sentiment, Orange-ambiguous).

It is worth noting that all three programmes, Pulsar, Caplena, and Infranodus, adhere to stringent privacy policies and ensure that no data is processed or sent outside of the EU. The NAP7 data uploaded to these AI tools will be retained solely for the duration of the current project and will be deleted six months after the project’s conclusion. Importantly, the data utilised in these programmes is strictly confined to the purposes of this research and is not employed for any other use. The programmes were also all used within UCL’s servers, which provide another level of security and protection.

## Results

In total, 10,746 responses to the survey were received, representing a 71% response rate [1]. Twenty-seven of the 57 survey questions required a free-text response. A final selection of 5,196 responses were then chosen to be analysed within the main themes of the initial paper [2].

Initially, for each of the key questions from the survey, researchers produced tables containing the themes derived from Infranodus, categorising the responses into positive or good experiences, negative or bad experiences, and neutral experiences. As discussed in the methodology, in the second stage of analysis, the human annotators identified the necessity to introduce an ’ambiguous’ category to capture the nuances in the free-text responses.

### Human vs Computer assisted analysis

Computer assisted sentiment tagged 36% (n=1871) of all 5196 themed responses as neutral, 20% (n=1039) as Negative, and 44% (n=2286) as positive. After nuances in some more complex answers initially rated as neutral by computer assisted tagging were spotted by human annotators, a second round of annotation with the newly developed sentiment framework (Fig 2) showed that far fewer comments fell within the context of neutral sentiment – 7% (n=406) versus the initial 36% (n=1871). Validity checking by human annotators found a much more even spread between ambiguous (n=355) and neutral sentiment (n=406) ∼ 7% each, while an extra 3% of responses were tagged as positive (47%, n= 2426), and 39% (n=2009) were tagged by humans as negative. The human-only analysis took 255 hours (Table 5- S3 Appendix), 210 hours (over about 26 days) of which consisted of coding the themes, while the machine-assisted coding only required 5 hours (Table 6- S3 Appendix). The computer-assisted analysis demonstrated a good range of accuracy of 72% in sentiment classification and 78% in thematic analysis, compared to the human-annotated dataset [2]. Inter-annotator agreement between human experts was 82%, indicating a good level of consistency in the manual annotation process [2]. However, the machine-assisted analysis required human intervention to refine the sentiment categories (40 hours), particularly in cases where the responses contained ambiguous or mixed sentiments, resulting in the machine-assisted analysis taking 88 hours in total. In Fig 2 below, the sentiment analysis framework shows the definition of each sentiment coded, including the differences between neutral and ambiguous within the context of this study.

The comparison of human and machine-assisted analysis approaches revealed several key findings. The machine-assisted analysis significantly reduced the time required to analyse the large dataset, enabling a more efficient processing of the qualitative data. While the overall percentages of positive and negative sentiments did not vary drastically between the computer-assisted tagging and human annotation, the introduction of the “ambiguous” sentiment category in the new framework allowed for a more nuanced understanding of the responses and insights into peri-operative cardiac arrest experiences. By distinguishing between truly neutral sentiments and those that were more complex or ambiguous, the human annotators were able to provide a more accurate representation of the respondents’ experiences.

### Thematic analysis

In terms of thematic analysis - this paper details the Big Qual methods by analysing themes from Tables 1-4, focusing on guidelines for managing peri-operative cardiac arrests, recent incident management, and debriefing processes [2]. Over six months, four researchers (EK, SM1, SM2, ND) analysed and categorised the NAP7 free-text data for 13 key questions, supporting the AI tools during the machine learning phase.

Table 1 below is an example output from the AI tools for key questions that helped researchers analyse the themes. Table 1 presents main themes such as Patient Outcome, Leadership and Teamwork, Management Procedures, Recognition of Arrest and Treatment, and Chaos. Additional themes (detailed in Tables 2-4 in the supplementary information S3 Appendix) include Awareness and Adequacy of Guidelines, Specific Scenarios (Table 2), Debrief (Table 3), Support, Confidence, and Mental Health Impact, and Learned from Experience, Overall Improvement (Table 4). All themes were human-annotated for sentiment.

**Table 1.** ‘Free-text’ comments and themes from 1869 anaesthetists regarding the management of the peri-operative cardiac arrest that they most recently attended. The themes and sentiment categories (positive, neutral/mixed, negative) were determined by the ML tools. The sentiment category ‘Ambiguous’ was added by human annotators. CALS, Cardiac advanced life support; ODP, operating department practitioners; PPE, personal protective equipment; ROSC, return of spontaneous circulation.

| Themes (Q2 and Q30 - number of sentiments) | Examples |
| --- | --- |
| Patient outcome (n=1182)<br>• *Positive comments (n=964)*<br>• *Neutral comments (n=22)*<br>• *Ambiguous comments (n=33)*<br>• *Negative comments (n=163)* | *Positive examples*<br>'Successful outcome, no issues, concerns.'<br>'Good initial outcome. Although very stressful as significant uncertainty over the actual diagnosis.'<br>*Neutral examples*<br>'Corroborated by Serious Incident investigation.'<br>'Discharged from hospital.'<br>*Ambiguous examples*<br>'The QRH is useful but doesn't go through the nuances and most likely causes for specific patient cohorts.'<br>'There still some grey areas with regards to advice, for example DNACPR patients undergoing surgery.'<br>*Negative examples*<br>'CALS protocol followed, chest opened and ROSC but not that well led by ICU consultant'.<br>'ROSC was obtained after first cardiac arrest...delay in transferring patient to intensive care... Patient then had second cardiac arrest' |
| Leadership and teamwork (n=313)<br>• *Positive comments (n=285)*<br>• *Neutral comments (n=2)*<br>• *Ambiguous comments (n=3)*<br>• *Negative comments (n=23)* | *Positive Examples*<br>'Good teamwork. Got child back very quickly.'<br>'Theatre team worked very well together. All commented that it had felt like Sim training.'<br>*Neutral examples*<br>'This happened in radiology and resuscitation and senior anaesthesia teams had to be summoned.'<br>'Leading ITU consultant led arrest and outcome wouldn't have changed.'<br>*Ambiguous examples*<br>'Never exposed to this scenario in real life or simulation, complex situation with multiple additional and complex factors compared with a 'standard' ward arrest (different team with variable skill set and mix - team management more complex, requires management from anaesthetic and surgical |
|  | perspective, consideration of anaesthetic and surgical as well as patient factors as a cause etc.)’<br>‘I am confident in the management of cardiac arrests in general, intra-operative is an unexpected and complex situation with a procedure at varying degrees of completion or ability to easily pause and considerations of sterility etc. Therefore, the human factors and team working in this scenario are often significantly challenging and unfamiliar.’<br>*Negative examples*<br>‘No leader, consultant in list in disarray, others helping in slightly uncoordinated fashion, but shocks delivered and outcome good.’<br>‘Consultant refused to recognise patient had arrested, had to overrule him’ |
| Management procedures e.g., guidelines (n=202)<br>• *Positive comments (n=169)*<br>• *Neutral comments (n=6)*<br>• *Ambiguous comments (n=8)*<br>• *Negative comments (n=19)* | *Positive examples*<br>‘Well managed, major haemorrhage protocol already activated.’<br>‘Good prompt resuscitation of patient. We followed the guidelines to a high standard.’<br>*Neutral examples*<br>‘Standard arrest management’<br>‘I came in as support during the arrest - I can't comment on the management at the start of the case.’<br>*Ambiguous examples*<br>‘I feel as if I understand the appropriate management and protocols but have never been in this scenario so hesitate to say I am confident.’<br>‘There are times however where there may be tasks, I would have to do, for example, manage the airway, making it difficult to also lead the management!’<br>*Negative examples*<br>‘Management was hampered by difficulty in communication and obtaining equipment due to COVID and PPE.’<br>‘Mandatory to put out hospital cardiac arrest call. Medical team unfamiliar with IR suite and the procedure being undertaken. Also unfamiliar with anaesthesia and standard processes that were underway.’ |
| Recognition of arrest and treatment (n=90)<br>• *Positive comments (n=83)*<br>• *Neutral comments (n=1)* | *Positive examples*<br>‘Early identification of deteriorating patient and appropriate management, whole arrest team was present before the event.’<br>‘There was a prompt recognition of the cardiac arrest with a |
| • Ambiguous comments (n=0)<br>• Negative comments (n=6) | high index of suspicion as to the cause throughout.'<br>*Neutral example*<br>'It was a peri arrest situation.'<br>*Negative examples*<br>'Consultant refused to recognise patient had arrested, had to overrule him to get ODP to start chest compressions.'<br>'Not recognised early enough. Poor communication from surgeon who insisted it must be an airway problem.' |
| Chaos (n=82)<br>• Positive comments (n=19)<br>• Neutral comments (n=4)<br>• Ambiguous comments (n=5)<br>• Negative comments (n=54) | *Positive example*<br>'Very difficult case with multiple problems at the same time. We did the best we could!'<br>*Neutral example*<br>'Initially very busy/chaotic as 2222 call put out and medical team arrived. Then ICU consultant (Echo trained) arrived and took over lead.'<br>*Ambiguous examples*<br>'Chaotic at the beginning.'<br>'Clinically managed well, patient had good outcome. Felt chaotic as input from medical team with minimal experience in perioperative arrest.'<br>*Negative examples*<br>'Too many people giving orders, disorganised.'<br>'A bit chaotic as a lot of people and equipment in a small room.'<br>'Chaotic environment with different people trying to lead.'<br>'A lot of people involved, sometimes difficult to see what is being or has been done.' |

We then further analysed the themes in Infranodus. The theme “chaos” refers to the chaotic nature of the management of the arrest. We use this theme to further demonstrate an example of the nuances in the clinicians’ responses using Infranodus and Caplena (Table 1), where the negative, positive, neutral, and ambiguous examples of “chaos” varied depending on the complexity of each situation. In Fig 3, topic analysis conducted in Infranodus provides a visual thematic network graph of: ‘What does the impact of good & bad chaos look like?’ The topic graph shows several topical clusters, with the most prominent words and themes appearing larger in size and provides insights into the key themes and connections related to different forms of chaos in a peri-operative environment, based on the responses analysed. The central subtheme is “patient”, which is closely connected to various aspects of the chaotic situation, such as the arrest, team, and management. The closer the nodes are together, the more linked/connected the topics are to each other. Key topical clusters include Teamwork Quality, Cardiac Arrest Management, Resuscitation Algorithms, and Decision Making. Contextual examples highlight challenges such as lack of clear leadership, inexperienced team members, and conflicting instructions. The orange boxes representing ambiguous statements refer to the example statement on the left-hand side of the figure as well as the orange topic box on the right-hand side of the figure. Overall, the topic graph emphasises the importance of effective teamwork, clear communication, well-defined protocols, and decisive leadership in managing chaotic situations in the peri-operative environment to ensure positive patient outcomes.

## Discussion

The comparison of human and machine-assisted analysis approaches in this study highlights the potential of AI-assisted tools in patient safety research. By using the AI tools Caplena and Infranodus to analyse the free-text data from the NAP7 survey we have aimed to validate the potential of AI-assisted tools in patient safety research. Our findings demonstrate that machine-assisted analysis can significantly reduce the time and effort required to process large volumes of qualitative data, enabling researchers to quickly identify key themes and sentiments. However, the results also show the importance of human expertise in refining and interpreting machine-generated outputs. The human analysis revealed the need for an additional sentiment category, “ambiguous”, which captured the complexity and nuances of the peri-operative cardiac arrest experiences. The insights gained from the qualitative analysis of peri-operative cardiac arrest experiences highlight several areas where focused efforts and quality improvement initiatives can be implemented.

Sentiment analysis plays a crucial role in understanding the complex emotions and experiences of healthcare professionals involved in peri-operative cardiac arrest events. In our study, sentiment analysis led by AI was essential for capturing the wide range of emotions expressed in the free-text responses of anaesthetists. By delineating the nuances of sentiment, including the introduction of the “ambiguous” category, we were able to provide a more comprehensive and accurate representation of the respondents’ experiences. This nuanced approach not only enriches the current research by highlighting the emotional intricacies of patient safety incidents but also sets a foundation for future studies to adopt more sophisticated sentiment analysis techniques. The ability to identify and interpret subtle differences in sentiment can inform targeted interventions, support staff wellbeing, and ultimately enhance patient safety practices.

The “ambiguous” category was introduced to represent statements that expressed a mix of positive, negative, and neutral sentiments, or where the overall sentiment was unclear. This category differed from “neutral” in that neutral statements were largely factual or objective, without expressing strong emotions or opinions. In contrast, ambiguous statements often contained conflicting sentiments or nuanced perspectives that could not be easily classified as positive, negative, or neutral.

The inclusion of the “ambiguous” category in the sentiment analysis framework (Tables 1 – 4) allowed for a more accurate representation of the complex emotions and experiences expressed by healthcare professionals in relation to patient safety incidents. Our findings show the importance of considering the emotional complexity and nuance alongside sentiment analysis to gain a more comprehensive understanding of the challenges faced by healthcare professionals during peri-operative cardiac arrests. By examining the findings through the lens of emotional complexity, we can identify key areas for improvement to enhance patient safety. The introduction of the “ambiguous” category highlights the value of human expertise and domain knowledge in refining ML models for patient safety analysis, ultimately leading to more accurate and actionable insights. These findings also underscore the complexity of staff experiences and the need to address the identified challenges and support for staff wellbeing.

It is essential to highlight that while the previous NAP7 paper [2] thoroughly demonstrated the impact of peri-operative cardiac arrest on anaesthetic staff, the present study focuses on the methods used to analyse the co-related themes, sentiments, and experiences of anaesthetic staff working in peri- operative cardiac arrest management, and a comparison of human and machine-assisted analysis approaches. The co-related themes identified in our analysis, such as teamwork, communication, leadership, patient outcomes, and the debriefing process (Tables 1-4), all played key roles in understanding the extent and nature of the impact of peri-operative cardiac arrest on anaesthetic staff. By focusing on these themes and their interrelationships, our study complements the findings of the previous NAP7 paper and provides a more nuanced understanding of staff experiences and the factors that shape them.

Tables 1-4 highlight several relevant themes related to patient safety incidents and the management of peri-operative cardiac arrests. The sentiment analysis examples reveal nuanced insights into the complexities of these situations.

For example, regarding the theme Teamwork (Table 1): Our analysis revealed that effective teamwork, communication, and leadership were crucial factors in determining the success of peri-operative cardiac arrest management [21]. Sujan and colleagues [21] also emphasise the importance of considering these factors alongside AI use in healthcare. The identification of these factors in the analysis of patient safety free-text data is emphasised by Ede et al [22] in their synthesis of literature on teamwork and communication in escalating acute ward care. A flattened hierarchy and team functioning were key themes identified in the literature [23]. In our data, team functioning and teamwork were regular themes identified in many of the questions about the impact of peri-operative cardiac arrest (Table 1 “Leadership and teamwork”). Table 1’s ambiguous examples related to leadership and teamwork emphasise the unique challenges posed by peri-operative cardiac arrests, such as unfamiliar team dynamics, complex scenarios, and the need for effective communication and coordination between anaesthetic and surgical teams. Where teams worked well together, team relationships and communication were good, anaesthetists spoke more positively about the events, even if the overall outcome was not positive – ambiguous comments had a mixture of both positive and negative/neutral sentiment. These insights underscore the importance of training and simulation exercises that focus on enhancing non-technical skills in high-stress situations.

Regarding the theme on Guidelines (Table 2) which refers to the remarks about guidelines used to manage the peri-operative cardiac arrest, notably, the ambiguous examples shed light on the challenges faced by anaesthetists and the need for targeted interventions. In Table 2, the ambiguous examples related to guidelines underscore the importance of addressing specific scenarios and nuances in peri- operative cardiac arrest management. Comments such as “The QRH is useful but doesn’t go through the nuances and most likely causes for specific patient cohorts” and “There [sic] still some grey areas with regards to advice, for example DNACPR patients undergoing surgery” highlight the need for guidelines that provide guidance on handling complex situations and patient-specific factors.

In terms of the debrief theme which referred to the debriefing processes respondents had after the cardiac arrest (Table 3), sentiment and thematic analysis revealed positive, negative, neutral, and ambiguous sentiments about the debriefing process following peri-operative cardiac arrests. While some respondents expressed positive sentiments about the opportunity to discuss and analyse the event, others reported negative experiences, such as criticism or lack of proper training by those conducting debriefs. The ambiguous examples in Table 3, related to debriefing, reveal the importance of a well-structured and psychologically safe debriefing process. Comments like “It felt disorganised with no communication or debrief but I felt satisfied because all the necessary steps were followed, and patient received best possible care” highlight the need for a balance between adhering to protocols and fostering open communication during debriefs. These sentiments emphasise the importance of a well-structured, inclusive, and psychologically safe debriefing process. One way to address this would be to work on establishing a process that encourages the participation of all team members and provides opportunities for individual debriefing sessions to address personal concerns or emotional needs. By promoting positive sentiments and minimising negative ones, healthcare organisations can foster a culture of learning, support, and continuous improvement. Healthcare organisations can leverage these insights to develop staff support, such as training programs and simulation exercises, to enhance the non-technical skills of their staff and foster a culture of effective communication, teamwork, and decision-making.

The theme of Psychosocial Impact and Support (Table 4), which refers to general social support and well-being of the anaesthetists and how they were impacted by the cardiac arrest, encompassed a range of sentiments, with many ambiguous experiences reported. The ambiguous examples highlight the complex nature of these incidents and their emotional toll on anaesthetists. For instance, one respondent commented, “I suppose it ended well so I didn’t feel too bad. I’m no [sic] so sure about the junior members of the anaesthetic team,” indicating a sense of uncertainty and concern for the well-being of their colleagues. Negative experiences were also prevalent, with anaesthetists expressing a lack of support from their organisations and increased anxiety when dealing with similar cases in the future. Positive experiences were less common but highlighted the importance of access to robust psychological support services and informal support from colleagues.

The insights gained from Table 4 emphasise the need for healthcare organisations to focus on providing structured support systems, including access to psychological services, peer support networks, and formal debriefing processes. Additionally, the findings highlight the importance of fostering a culture of learning and continuous improvement, where anaesthetists can openly discuss their experiences, share lessons learned, and receive constructive feedback to enhance their practice.

Future research in this area could explore the application of our novel Big Qual methods and AI-assisted analysis tools to other healthcare settings and patient safety incidents, such as medication errors or surgical complications. Additionally, studies could investigate the effectiveness of targeted interventions based on the insights gained from this research, such as improved debriefing processes, scenario-specific guidelines, and team training programmes. Researchers could also focus on refining machine learning models to better capture the nuances and complexities of healthcare professionals’ experiences, ultimately leading to more accurate and actionable insights for enhancing patient safety. Furthermore, collaborations between healthcare professionals, data scientists, and patient safety experts could help develop standardised methodologies and best practices for applying AI-assisted qualitative analysis in healthcare, ensuring the responsible and ethical use of these powerful tools. By continuing to explore the potential of Big Qual methods and AI-assisted analysis in patient safety research, we can work towards creating safer healthcare environments and improving outcomes for both patients and healthcare professionals.

### Limitations

Our study has limitations that should be considered when interpreting the findings. First, while the overall response rate of 71% is similar to that of the NAP6 baseline survey [22], the response rates for each question progressively decreased from 100% to 92%, potentially introducing bias if non-respondents had different experiences or perspectives. However, the bias will be limited due to the whole population of anaesthetists being sampled, not just a representative sample within this population. Second, there is a risk of self-selection bias, as those with particularly memorable or impactful experiences might have been more inclined to participate, potentially skewing the results towards more extreme cases. Additionally, the specific focus on peri-operative cardiac arrests and the sole inclusion of anaesthetists’ perspectives may limit the generalisability of our findings to other healthcare settings, patient safety incidents, and the viewpoints of other resuscitation team members.

Despite our adherence to information governance and data anonymisation, there are potential challenges and ethical considerations associated with implementing AI-assisted qualitative analysis in healthcare settings. Previous studies have highlighted the need for human oversight and interpretation [10] which we have tried to mitigate by introducing human input at each stage. However, the possibility of algorithmic bias in the initial training data of the AI computer tools used still perpetuates AI health inequalities based on lack of diversity in that specific AI training data [24, 25].

Furthermore, although we had five researchers from different backgrounds and specialties involved in the thematic analysis and three during the sentiment analysis process, we only had two researchers in the discussions of the percentage agreement of the human analysis. These researchers will have brought their own subjectivity to the analysis. If there had been more team members, the percentage agreement may have been lower as there may have been more varied results across multiple researchers. Another potential limitation of our study is the involvement of the same human analysts in both the human-only and machine-assisted analyses. As the analysts gained familiarity with the data during the human-only analysis, there is a possibility of learning or spillover of knowledge when conducting the machine-assisted analysis. This familiarity could have influenced their interpretations and decisions during the machine-assisted analysis, potentially leading to a more refined or biased understanding of the data compared to analysts without prior exposure. However, the use of a structured sentiment analysis framework (Fig 2) and the involvement of three analysts from different backgrounds and specialties (EB, EK, SM1) at times, aimed to mitigate this potential bias by ensuring consistency and reducing individual subjectivity in the analysis process.

Despite these limitations, our study represents the analysis of the largest survey of anaesthetists to date examining individual preparedness, management, and experiences of peri-operative cardiac arrest. The use of Big Qual methods and an adaptive sentiment analysis and thematic analysis framework highlights the potential for applying these techniques to other areas of patient safety research. The insights gained can guide the development of tailored strategies and quality improvement initiatives to enhance patient safety.

## Conclusion

This study employed a novel approach using AI-assisted data analysis tools to examine a substantial body of free-text survey data. The primary focus was to compare the effectiveness of machine-assisted analysis, using tools like Infranodus and Caplena, against human-only analysis in identifying themes, sentiments, and insights. The free-text responses from the Royal College of Anaesthetists’ 7th National Audit Project (NAP7) baseline survey on peri-operative cardiac arrest experiences served as a case study to test and validate this approach.

The machine-assisted analysis significantly streamlined the process of analysing the large dataset, enabling the efficient identification of key themes and sentiments. While the overall percentages of positive and negative sentiments did not differ drastically between the computer-assisted tagging and human annotation, the human input played a crucial role in introducing the “ambiguous” sentiment category. This new category allowed for the capture of more complex and nuanced responses that were initially tagged as neutral by the computer. By distinguishing between truly neutral sentiments and those that were more ambiguous, the human annotators provided a more comprehensive understanding of the anaesthetists’ experiences, highlighting the value of human expertise in refining the sentiment analysis.

The insights gained from this study also complement the findings of the previous NAP7 paper [2] by providing a more in-depth understanding of the Big Qual methodology employed to investigate staff experiences.

The application of AI-assisted data analysis tools, combined with human expertise, offers a powerful approach to efficiently analyse large-scale qualitative datasets while preserving the nuance and complexity of the data. This study demonstrates the potential of this novel methodology to streamline the analysis process, reduce resource requirements, and generate meaningful insights from Big Qual data. The integration of NLP, ML, and human input allows for a more comprehensive understanding of the themes, sentiments, and experiences captured in free-text responses. As AI technologies continue to advance, their application in analysing diverse sources of qualitative data can add further value to the field of qualitative research, enabling researchers to uncover valuable insights and inform data-driven decision-making across various domains. This study underscores the importance of continued interdisciplinary collaboration among domain experts, data scientists, and AI specialists to refine and optimise these methods, ensuring their reliability, validity, and ethical application in real-world contexts.

## Supporting information

NAP7 Survey

Collaborator list

## Data Availability

All data for our study is based on secondary data analysis of the data shared in the data available in online Supporting Information from the following publication Kane et al (2022) Methods of the 7th National Audit Project (NAP7) of the Royal College of Anaesthetists: peri-operative cardiac arrest. Anaesthesia 77: 1376-1385.

## Acknowledgements

The NAP7 panel and team members (collaborators) are listed in the supporting information (S1). We thank all NAP7 Local reporters and their teams and all UK anaesthetists who completed surveys. We also thank Dr Paulina Bondaronek for her feedback on the methods section of the paper.

## Supporting information captions

S1 Appendix – Collaborator list

S2 Appendix-NAP7 Baseline Survey

S3 Appendix -Tables 2-6

## References

1. Kane AD, Armstrong RA, Kursumovic E, Cook TM, Oglesby FC, Cortes L. et al. Methods of the 7th National Audit Project (NAP7) of the Royal College of Anaesthetists: peri-operative cardiac arrest. Anaesthesia. 2022 Dec;77(12):1376–85. 10.1111/anae.15856.

2. Kursumovic E, Cook TM, Lucas DN, Davies MT, Martin S, Kane AD. Et al. The 7th National Audit Project (NAP7) baseline survey of individual anaesthetists: preparedness for and experiences of peri-operative cardiac arrest. Anaesthesia. 2023 Dec;78(12):1453–64. 10.1111/anae.16154.

3. Martin S, Clark SE, Gerrand C, Gilchrist K, Lawal M, Maio L. et al. Patients’ experiences of a sarcoma diagnosis: a process mapping exercise of diagnostic pathways. Cancers. 2023 Aug 3;15(15):3946. 10.3390/cancers15153946.

4. Brower RL, Jones TB, Osborne-Lampkin LT, Hu S, Park-Gaghan TJ. Big qual: Defining and debating qualitative inquiry for large data sets. International Journal of Qualitative Methods. 2019 Nov 15;18. 10.1177/1609406919880692.

5. World Health Organization. Ethics and governance of artificial intelligence for health: guidance on large multi-modal models, 2024, [cited 5 February 2024], available from: https://iris.who.int/bitstream/handle/10665/375579/9789240084759-eng.pdf?sequence=1.6.

6. Karafillakis E, Martin S, Simas C, Olsson K, Takacs J, Dada S. et al. Methods for social media monitoring related to vaccination: systematic scoping review. JMIR public health and surveillance. 2021 Feb 8;7(2):e17149. http://publichealth.jmir.org/2021/2/e17149/.

7. Bates DW, Levine D, Syrowatka A, Kuznetsova M, Craig KJ, Rui A. et al. The potential of artificial intelligence to improve patient safety: a scoping review. NPJ digital medicine. 2021 Mar 19;4(1):54. 10.1038/s41746-021-00423-6.

8. Martin S, Kilich E, Dada S, Kummervold PE, Denny C, Paterson P. et al. “Vaccines for pregnant women…?! Absurd”–Mapping maternal vaccination discourse and stance on social media over six months. Vaccine. 2020 Sep 29;38(42):6627–37. 10.1016/j.vaccine.2020.07.072.

9. Guetterman TC, Chang T, DeJonckheere M, Basu T, Scruggs E, Vydiswaran VV. Augmenting qualitative text analysis with natural language processing: methodological study. Journal of medical Internet research. 2018 Jun 29;20(6):e231. https://www.jmir.org/2018/6/e231.

10. Towler L, Bondaronek P, Papakonstantinou T, Amlôt R, Chadborn T, Ainsworth B. et al. Applying machine-learning to rapidly analyze large qualitative text datasets to inform the COVID-19 pandemic response: comparing human and machine-assisted topic analysis techniques. Frontiers in Public Health. 2023 Oct 31;11:1268223. 10.3389/fpubh.2023.1268223.

11. Evans HP, Anastasiou A, Edwards A, Hibbert P, Makeham M, Luz S. et al. Automated classification of primary care patient safety incident report content and severity using supervised machine learning (ML) approaches. Health informatics journal. 2020 Dec;26(4):3123–39. 10.1177/14604582198331.

12. Infranodus Software [cited 29 September 2023]. Available from: https://infranodus.com/.

13. Paranyushkin D. Infranodus: Generating insight using text network analysis. InThe world wide web conference 2019 May 13 (pp. 3584-3589). 10.1145/3308558.3314123.

14. Caplena v2 (Caplena AG, Zurich, Switzerland), [cited 1 February 2024]. Available from: https://caplena.com/en/about/.

15. Momentive. SurveyMonkey. Niskayuna, NY: Momentive, 2023. [cited 2 February 2024]. Available from: SurveyMonkey - Free online survey software and questionnaire tool.

16. Pulsar Platform v2022 (Pulsar TRAC, first-party data tool, Pulsar Platform, London, UK). [Cited 1 November 2023]. Available from: https://www.pulsarplatform.com/solutions/audience-insights.

17. O’Brien BC, Harris IB, Beckman TJ, Reed DA, Cook DA. Standards for reporting qualitative research: a synthesis of recommendations. Academic medicine. 2014 Sep 1;89(9):1245–51. 10.1097/ACM.0000000000000388.

18. Hutto C, Gilbert E. Vader: A parsimonious rule-based model for sentiment analysis of social media text. In Proceedings of the international AAAI conference on web and social media 2014 May 16 (Vol. 8, No. 1, pp. 216–225). 10.1609/icwsm.v8i1.14550.

19. McHugh ML. Interrater reliability: the kappa statistic. Biochemia medica. 2012 Oct 15;22(3):276–82. https://hrcak.srce.hr/89395.

20. Kelly FE, Frerk C, Bailey CR, Cook TM, Ferguson K, Flin R. et al. Human factors in anaesthesia: a narrative review. Anaesthesia. 2023 Apr;78(4):479–90. 10.1111/anae.15920.

21. Sujan MA, Pozzi S, Valbonesi C. Reporting and learning: from extraordinary to ordinary. In Resilient Health Care, Volume 3 2016 Oct 3 (pp. 103–110). CRC Press.

22. Ede J, Petrinic T, Westgate V, Darbyshire J, Endacott R, Watkinson PJ. Human factors in escalating acute ward care: a qualitative evidence synthesis. BMJ open quality. 2021 Feb 1;10(1):e001145. 10.1136/bmjoq-2020-001145.

23. Kemp HI, Cook TM, Thomas M, Harper NJ. UK anaesthetists’ perspectives and experiences of severe perioperative anaphylaxis: NAP6 baseline survey. BJA: British Journal of Anaesthesia. 2017 Jul 1;119(1):132–9. 10.1093/bja/aex124.

24. Leslie D, Mazumder A, Peppin A, Wolters MK, Hagerty A. Does “AI” stand for augmenting inequality in the era of covid-19 healthcare? BMJ. 2021 Mar 16;372. 10.1136/bmj.n304.

25. Agarwal R, Bjarnadottir M, Rhue L, Dugas M, Crowley K, Clark J. et al. Addressing algorithmic bias and the perpetuation of health inequities: An AI bias aware framework. Health Policy and Technology. 2023 Mar 1;12(1):100702. 10.1016/j.hlpt.2022.100702.

