## Supplementary material for "Comparing human vs. machine-assisted analysis to develop a new approach for Big Qualitative Data Analysis": Tables 2-6

**Appendix S3 Tables 2-6**

**Table 2. Underlying themes from ‘free text’ comments (*n* = 2754) on respondents reporting on the question ‘Are existing guidelines for the management of peri-operative cardiac arrest sufficient? (Please provide reasons for your answer)’.** The themes and sentiment categories (positive, neutral, negative) were determined by the ML tools. The sentiment category ‘ambiguous’ was added by human annotators. Comments from one respondent may have created one or more themes. *AAGBI, Association of Anaesthetists of Great Britain and Ireland; ALS, Advanced Life Support; DNACPR, Do Not Attempt Cardiopulmonary Resuscitation; QRH, Quick Reference Handbook; RCUK, Resuscitation Council (UK).*

| **Themes (Q4 - number of sentiments)** | **Examples** |
| --- | --- |
| Awareness of guidelines (*n* = 1002):   - Positive comments (*n* = 239) - Neutral comments (*n* = 86) - Ambiguous comments (n = 44) - Negative comments (*n* = 633) | *Positive examples*  ‘AAGBI QRH provides a guide which is more tailored to the peri-operative cardiac arrest, compared with ALS.’  ‘Familiar. Generally easy to follow in high pressures arrest situation.’  ‘ALS guidelines offer good evidence-based algorithms.’  ‘We are following national and international guidelines which are created by the most experience colleagues in the management of cardiac arrest.’  ‘Training available and guidelines are readily available too.’  ‘AAGBI quick reference guidelines are pretty good.’    *Neutral examples*  *‘*Most are based on medical reasoning’  'Complexity of cases sometimes demands lateral thinking'  ‘This is most commonly a special circumstance'  ‘Very niche area'  *Ambiguous examples*  'Although I have had training in resuscitation fairly recently, I feel the theatre environment is unique and needs special guidelines with frequent updates'  ‘Are there any?’  'Although comfortable with ALS guidelines and theatre setting, would be nice to have training on specific theatre scenarios e.g. prone/open abdomen'    *Negative examples*  ‘I have not recently read these guidelines.’  ‘I have not delved into them in much detail.’  ‘Not aware of specific peri-operative guidelines.’  ‘I do not know where to access them or what the existing guidelines are.’  ‘No one seems to know the guidelines. Arrest teams are called by junior team members when not needed.’  ‘I didn't know there was a guideline!’  ‘I'm not aware of any formal guidelines for intraoperative arrest specifically.’ |
| Adequate guidelines (*n* = 1219):   - Positive comments (*n* = 383) - Neutral comments (*n* =159) - Ambiguous comments (n = 66) - Negative comments (*n* = 611) | *Positive examples*  ‘The guidelines provide clear information on the management of peri-operative cardiac arrests.’  ‘Baseline algorithm is sound, and guidelines need to be concise enough to act as quick reference and training aid.’  ‘We have ALS guidelines at hand in the event of peri-operative arrest that are clear, concise and easy to follow.’  ‘The QRH is very thorough and good to have as an app on my phone, plus available in all anaesthetic rooms.’  ‘Written guidelines and crisis cards are readily available to guide management.’    *Neutral examples*  ‘Can be more visible in ALS’  ‘I think it may be better if we can have training in own theatre setting with own staff'  ‘Peri operative situations are so varied and vast that I don't know if it is possible to have a comprehensive guideline on this'    *Ambiguous examples*  'There are various guidelines for the management of emergencies e.g. anaphylaxis, cardiac arrest. However, these do not account for situations specific to surgery e.g. surgical tools / open abdomen or chest.’  'Guidelines can always be improved/updated. But not clear to me if significant changes have occurred. Having said that specific scenarios with newer surgical technology probably require attention.’  'I'm not sure how easy they are to apply in real life'  'Guideline for emergency situations present but arrest algorithms generic, unsure is specific perioperative arrest guidelines would be beneficial'    *Negative examples*  ‘Needs to include more on team roles.’  ‘As above – RCUK is really focused on non-theatre arrests – see recent editorial on challenging 'no trace wrong place' for example!’  ‘Need clarity for specific situations including where respect forms are completed and DNACPR instituted.’ |
| Specific scenarios (*n* = 533)   - Positive comments (*n* = 58) - Neutral comments (*n* = 22) - Ambiguous comments (n = 63) - Negative comments (*n* = 390) | *Positive examples*  ‘Our scenario based, in theatre training (for consultants, with consultants) is excellent.’  ‘Plenty of info available for peri-operative deterioration, cardiac arrest and management.’  *Neutral examples:*  ‘The bare bones of guidelines are fine but the nuances of a perioperative arrest could perhaps be served well with specific guidance’  ‘RCOA guidelines seem to just point to RCUK guidelines for adults and children, not specific for intraoperative’    *Ambiguous examples*  ‘The caveat is whether to proceed or cancel surgery'  ‘Awareness that intra operative management has its own specific guidelines'  'Include operative causes of cardiac arrest e.g. intralipid'  ’ You can't be too didactic with these nuanced scenarios ‘  ‘Guidelines can always be improved/updated. But not clear to me if significant changes have occurred. Having said that specific scenarios with newer surgical technology probably require attention.’    *Negative examples*  ‘Peri-operative cardiac arrest differs from other in hospital arrests and needs to be treated as a special situation.’  ‘Doesn't always take into account different team structure (e.g. no medics, anaesthetic lead, theatre team).’  ‘This does not mention about some scenarios like when patient is in prone position or having surgery in head and neck area where table is turned away from anaesthetic machine. It needs some training in terms of ergonomics or logistics.’ |

**Table 3.** **Underlying themes from ‘free text’ comments (*n* = 313) on respondents reporting on the question ‘I was satisfied with the debrief process following the event’.** The themes and sentiment categories (positive, neutral, negative) were determined by the ML tools. The sentiment category ‘Ambiguous’ was added by human annotators. Comments from one respondent may have created one or more themes. *MDT, multidisciplinary team.*

| **Themes (Q36 - number of sentiments)** | **Examples** |
| --- | --- |
| Debrief (n= 313)   - Positive experience (*n* = 194) - Neutral experience (*n* = 22) - Ambiguous experience (n= 21) - Negative experience (*n* = 76) | *Positive examples*  ‘Everyone at the arrest were present. All contributed. Those that had seemed shaken at the event, looked happier after the debrief.’  ‘Everyone had the chance to speak and analyse the events leading up to the airway loss during tracheostomy insertion.’  ‘Informal debrief was satisfactory to all, in view of positive outcome. Team all well known to one another and able to talk openly and supportively.’  *Neutral examples:*  ‘Informal led by a surgeon not trained in debriefing. Would have benefited from a further cold debrief.’  ‘Would have been good to do a cold debrief with MDT but difficult due to shift work.’  *Ambiguous examples*  ‘It felt disorganised with no communication or debrief but I felt satisfied because all the necessary steps were followed, and patient received best possible care."  ‘a lot about looking after team in principle and debrief but not about talking to relatives or actual how’  *Negative examples*  ‘The whole process was so traumatising. On reflection, I feel we need two types of formal debriefs - hot and cold.’  ‘Was conducted in the wrong way for a hot debrief and led to a lot of upset and feelings of criticism.’  ‘It involved anyone involved in the arrest so difficult for consultant anaesthetists to open up with very junior members of the team there. Also didn't really discuss what went well, what could be improved. No individual debriefing occurred.’ |

**Table 4.** **Thematic analyses of anaesthetists reporting impact on future patient care delivery following most recent peri-operative cardiac arrest (n = 260).** The themes and sentiment categories (positive, neutral, negative) were determined by the ML tools. The sentiment category ‘Ambiguous’ was added by human annotators. Comments from one respondent may have created one or more themes.

| **Themes (Q41 and 43 - number of sentiments)** | **Examples** |
| --- | --- |
| Psychosocial impact and support (n= 129)   - Positive experience (*n* = 12) - Neutral experience (*n* = 26) - Ambiguous experience (n= 61) - Negative experience (*n* = 30) | *Positive examples*  ‘I sourced my own help on the advice of a consultant colleague. This was through NHS Practitioner Health (psychological support) and the service provided was excellent.’  ‘Excellent informal support from consultant involved’  ‘Good service in those needing psychological assistance’  *Neutral examples:*  ‘Wellbeing hospital support available but I have not felt the need to access’  ‘Didn't need more support’  *Ambiguous examples*  ‘I suppose it ended well so I did feel too bad. I'm no [sic] so sure about the junior members of the anaesthetic team’  ‘Noticed much more caution/anxiety/awareness around giving drugs. Not necessarily a bad thing.' 'I am definitely very very prepared for this now! Increased the stress I think after seeing what my colleague went through on that day’  *Negative examples*  ‘Support, if any, is from colleagues. Nothing from the organisation. Medical staff are simply expected to brush such events off, and get on with the next case’  ‘More cautious/ anxious about anaesthetising undiagnosed metabolic children’ |
| Learned from experience, improvement overall (n= 131)   - Positive experience (*n* = 22) - Neutral experience (*n* = 6) - Ambiguous experience (n= 51) - Negative experience (*n* = 52) | *Positive examples*  ‘Made me a bit more confident in dealing with this type of emergency.' 'I had recently completed ALS’  ‘Made me more cautious - made me better at delivering care. Improved my consenting’  *Neutral examples:*  ‘We have recently introduced peer support but I have no felt the need to access it following this event’  *Ambiguous examples*  ‘Informal discussions had but "needed" by myself. I came to help. Harder when you are responsible before arrest.’  *Negative examples*  ‘loss of confidence and questioning of my competence’  ‘I took days off, didn't want to look after patients’ |

**Table 5. Human-only analysis procedure and person-hours**

| Step in process | Procedure | Hours |
| --- | --- | --- |
| Preparation | Preparation of data from the survey. Data cleaning and extraction into Excel spreadsheets for manual sentiment analysis checking | 25 |
| Coding | - Manual sentiment-analysis -coding of sentiment for all sub-themes to come from InfraNodus and Caplena. - Thematic analysis by hand | - 100 (3 researchers)      - 110 (4 researchers) |
| Validity checks | Cross-checked 4 out of 13 questions (30%) for sentiment analysis | 15 |
| Interpretation | Interpreting the sentiment analysis and writing up | 5 |
| Total person hours | | 255 |

**Table 6.** **Machine-assisted analysis procedure and person-hours**

| Step in process | Procedure | Hours |
| --- | --- | --- |
| Preparation | Data cleaning from survey and importing CSV files into Caplena and InfraNodus | 30 (15 hours per analysis tool) |
| Coding | The computing of themes and sentiments of each question in both programmes | 5 |
| Validity checks/ training the analysis tool | Caplena - checking of themes/sub-themes (re-labelling) | 40 |
| Interpretation | Thematic and sentiment analysis written up from InfraNodus and Caplena outputs | 13 (1 hour per question) |
| Total person hours | | 88 |
