## Supplementary material for "Comparing human vs. machine-assisted analysis to develop a new approach for Big Qualitative Data Analysis": NAP7 Survey

### Baseline survey of all anaesthetists

#### 1. Welcome to the NAP7 Baseline Survey: Anaesthetists and Anaesthesia Associates

**Many thanks for taking part in this first phase of NAP7: Perioperative Cardiac Arrest in the UK.**

**This survey will take 5-10 minutes to complete. Please complete in one sitting, otherwise your data may be lost.**

**NAP7 will start on 16th June 2021. Please report cases of perioperative cardiac arrest in adults and children to your Local Coordinator.**

#### 2. Welcome to the NAP7 Baseline Survey: Anaesthetists and Anaesthesia Associates

**Some questions viewed through NHS browsers may not appear correctly. You may wish to use your own device.**

**SCOPE: This survey is about perioperative cardiac arrest including your attitudes and experience, training and demographics.**

**METHODS: This survey is for all anaesthetists and all anaesthesia associates working in all UK hospitals (NHS and Independent sector). All grades, including trainees, should complete this survey.**

**ALL responses are confidential and anonymous.**

#### 3. Knowledge, training and attitudes of perioperative cardiac arrest

* 1. When was your **most recent** training in advanced life support including chest compressions and defibrillation? Tick one option for each row.

*Resuscitation Council UK (RCUK) or equivalent courses:* *e.g. ALS, ILS, APLS, EPALS*

*In-house hands-on training sessions:* *departmental or hospital*

| | Instruct at least yearly | Within 1 - 2 years | Within 2 - 4 years | > 4 years | Not applicable (e.g. do not treat children) | None | Can't recall |
|---|---|---|---|---|---|---|---|
| RCUK adult or equivalent | ◯ | ◯ | ◯ | ◯ | ◯ | ◯ | ◯ |
| In-house adult 'hands-on training' | ◯ | ◯ | ◯ | ◯ | ◯ | ◯ | ◯ |
| RCUK paediatric or equivalent | ◯ | ◯ | ◯ | ◯ | ◯ | ◯ | ◯ |
| In-house paediatric 'hands-on training' | ◯ | ◯ | ◯ | ◯ | ◯ | ◯ | ◯ |

* 2. Do you agree or disagree with the following statement?

**I am confident in leading the management of cardiac arrest on the operating table.**

| Strongly agree | Agree | Neither agree or disagree | Disagree | Strongly disagree |
|---|---|---|---|---|
| ◯ | ◯ | ◯ | ◯ | ◯ |

Please provide reason(s) for your answer.

* 3. Do you agree or disagree with the following statements?

| | Strongly agree | Agree | Neither agree or disagree | Disagree | Strongly disagree |
|---|---|---|---|---|---|
| I have received sufficient training in the management of **intraoperative** cardiac arrest. | ◯ | ◯ | ◯ | ◯ | ◯ |
| I would benefit from more training in the management of **intraoperative** cardiac arrest. | ◯ | ◯ | ◯ | ◯ | ◯ |
| I am confident in leading a debrief process. | ◯ | ◯ | ◯ | ◯ | ◯ |
| I would benefit from training in how to conduct a debrief. | ◯ | ◯ | ◯ | ◯ | ◯ |
| I am confident in leading communication with relatives/next of kin after an intraoperative cardiac arrest. | ◯ | ◯ | ◯ | ◯ | ◯ |

* 4. Do you agree or disagree with the following statement?

**Existing guidelines for the management of perioperative cardiac arrest are sufficient.**

| Strongly agree | Agree | Neither agree or disagree | Disagree | Strongly disagree |
|---|---|---|---|---|
| ◯ | ◯ | ◯ | ◯ | ◯ |

Please provide reason(s) for your answer.

* 5. In your opinion, what are the **three most common** causes of perioperative cardiac arrest?

State 'not sure' if you are unsure.

Cause 1

Cause 2

Cause 3

* 6. In an anaesthetised 50-year old ASA 2 patient, without an arterial line, who developed hypotension, **whilst treating causes of profound hypotension**, what would you use as an indication to start chest compressions? Tick all that apply.

*(for BP, please choose the highest BP option at which you would start chest compressions)*

- [ ] Systolic BP 51 – 60 mmHg
- [ ] Systolic BP 41 – 50 mmHg
- [ ] Systolic BP 31 – 40 mmHg
- [ ] Systolic BP ≤ 30 mmHg
- [ ] Other (please specify)
- [ ] Unrecordable BP
- [ ] No palpable peripheral pulse
- [ ] No palpable central pulse
- [ ] Very low end-tidal CO2
- [ ] None of these
- [ ] I'm not sure

* 7. What indications would you use to start chest compressions for the previous question (Q6) if the patient was aged 75, hypertensive and ASA 3? Tick all that apply.

*(for BP, please choose the highest BP option at which you would start chest compressions)*

- [ ] Systolic BP 51 – 60 mmHg
- [ ] Systolic BP 41 – 50 mmHg
- [ ] Systolic BP 31 – 40 mmHg
- [ ] Systolic BP ≤ 30mmHg
- [ ] Other (please specify)
- [ ] Unrecordable BP
- [ ] No peripheral pulse
- [ ] No central pulse
- [ ] Very low end tidal CO2
- [ ] None of these
- [ ] I'm not sure

#### 4. Personal experience of perioperative cardiac arrest

**The following questions are about your recent experience (within the last 2 years).**

**Cardiac arrest is defined as the need for at least 5 chest compressions and/or defibrillation in a patient having a procedure under the care of an anaesthetist.**

**This question is for arrests occurring between your first hands-on contact with the patient at the start of anaesthesia care until handover to another clinician (e.g. leaving recovery area to the ward, handover to ICU).**

**PLEASE EXCLUDE cases where:**
**1. defibrillation is a planned, normal, or expected part of the procedure (e.g. during VT ablation);**
**2. chest/internal cardiac compressions and/or defibrillation occur during cardiopulmonary bypass;**
**3. patients in whom chest compressions and/or defibrillation were not started when cardiac arrest occurred;**
**4. patients who received <5 chest compressions.**

* 8. Within the **last 2 years**, how many cases of perioperative cardiac arrest do you recall being involved with (**present during or managed**)?

- ◯ 0
- ◯ 1
- ◯ 2
- ◯ 3
- ◯ 4
- ◯ 5
- ◯ More than 5
- ◯ Can't recall

#### 5. Most recent experience of perioperative cardiac arrest

**The following questions are about your most recent perioperative cardiac arrest in which you were involved (present during or managed).**

* 9. What was the location of the perioperative cardiac arrest?

* 10. What PPE precautions did you use during the management of the perioperative cardiac arrest?

**Airborne** = FFP3, fluid repellent long sleeved gown, gloves, eye protection
**Droplet** = Fluid resistant surgical mask, apron, gloves +/- eye protection
**Contact** = Standard face mask, apron, gloves +/- eye protection
**Other** = Standard face mask +/- gloves or no PPE

| | Airborne precautions | Droplet precautions | Contact precautions | Other | Can't recall |
|---|---|---|---|---|---|
| Just before/ at the time of arrest | ◯ | ◯ | ◯ | ◯ | ◯ |
| During the arrest | ◯ | ◯ | ◯ | ◯ | ◯ |

* 11. How would you describe your experience in managing a perioperative cardiac arrest **in PPE** compared to before the COVID-19 pandemic?

| Much worse | Worse | Neither better or worse | Better | Much better | Not applicable |
|---|---|---|---|---|---|
| ◯ | ◯ | ◯ | ◯ | ◯ | ◯ |

Please provide reason(s) for your answer.

* 12. What was the approximate age of the patient?

- ≤1 years old
- >1 – 5 years old
- >5 – 12 years old
- >12 – 18 years old
- >18 – 65 years old
- >65 years old
- Age not known
- Can't recall
- Prefer not to say

#### 6. Most recent experience of perioperative cardiac arrest

* 13. What was the most likely **‘suspected or confirmed’** primary cause of the perioperative cardiac arrest?

- Airway/breathing problem
- Cardiac/cardiovascular (including arrhythmia, MI, bleeding, sepsis, thromboembolic)
- Neurological problem
- Regional anaesthesia (including high neuraxial block, LA toxicity)
- Metabolic (including electrolyte disorder, and malignant hyperthermia)
- Anaphylaxis
- Error, drug or equipment problem
- Uncertain cause - multiple comorbidities and/or extreme age or frailty
- Unknown cause
- Can't recall

#### 7. Most recent experience of perioperative cardiac arrest

* 14. What was the most likely **‘suspected or confirmed’** primary cause of the perioperative cardiac arrest?

Choose single best option.

*If option not available, press 'previous' below for another category OR choose 'other' and state below.*

- ◯ Failed mask ventilation
- ◯ Failed supraglottic airway placement
- ◯ Failed intubation
- ◯ Laryngospasm
- ◯ Cannot intubate cannot oxygenate (CICO)
- ◯ Unrecognised oesophageal intubation
- ◯ Airway haemorrhage
- ◯ Regurgitation
- ◯ Aspiration of gastric contents
- ◯ Aspiration of blood
- ◯ Severe hypoxaemia
- ◯ Bronchospasm
- ◯ High airway pressure / obstructive ventilation
- ◯ Gas trapping / high iPEEP
- ◯ Hypercapnia
- ◯ Hypocapnia
- ◯ Pneumothorax
- ◯ Tension pneumothorax
- ◯ Endobronchial intubation
- ◯ Pulmonary embolism
- ◯ Ventilator disconnection
- ◯ Wrong gas supplied/unintentional connection to air
- ◯ Other (please specify)

#### 8. Most recent experience of perioperative cardiac arrest

* 15. What was the most likely **PRIMARY** cause of the perioperative cardiac arrest?

Choose single best option.

*If option not available, press 'previous' below for another category OR choose 'other' below.*

- ◯ Major haemorrhage
- ◯ Bradyarrhythmia
- ◯ Tachyarrhythmia
- ◯ Isolated severe hypotension (central vasopressors considered/started)
- ◯ DC cardioversion
- ◯ Cardiac ischaemia
- ◯ Cardiac tamponade
- ◯ New AF
- ◯ Ventricular tachycardia
- ◯ Ventricular fibrillation
- ◯ Complete heart block
- ◯ Pulmonary embolism
- ◯ Fat embolism
- ◯ Bone cement implantation syndrome
- ◯ Amniotic fluid embolism
- ◯ Air embolism
- ◯ $CO_2$ embolism
- ◯ Septic shock
- ◯ Anaphylaxis
- ◯ Local anaesthetic toxicity (excessive dose and/or wrong route)
- ◯ Incompatible blood transfusion
- ◯ Addisonian crisis
- ◯ Vagal outflow – e.g. pneumoperitoneum, oculo-cardiac reflex
- ◯ High neuraxial block
- ◯ Other (please specify)

#### 9. Most recent experience of perioperative cardiac arrest

* 16. What was the most likely **PRIMARY** cause of the perioperative cardiac arrest?

Choose single best option.

*If option not available, press 'previous' below for another category OR choose 'other' below.*

- Intracranial haemorrhage (including subarachnoid haemorrhage)
- Raised intracranial pressure (e.g. new fixed/dilated pupil(s))
- Seizure
- Vagal outflow – e.g. pneumoperitoneum, oculo-cardiac reflex
- High neuraxial block
- Cushing's Response/Coning
- Neurogenic/spinal shock
- Stroke
- Local anaesthetic toxicity (excessive dose and/or wrong route)
- Other (please specify)

#### 10. Most recent experience of perioperative cardiac arrest

* 17. What was the most likely **PRIMARY** cause of the perioperative cardiac arrest?

Choose single best option.

*If option not available, press 'previous' below for another category OR choose 'other' below.*

- High neuraxial block
- Inadvertent neuraxial block during regional block
- Local anaesthetic toxicity (excessive dose and/or wrong route)
- Other (please specify)

#### 11. Most recent experience of perioperative cardiac arrest

* 18. What was the most likely **PRIMARY** cause of the perioperative cardiac arrest?

Choose single best option.

*If option not available, press 'previous' below for another category OR choose 'other' below.*

- New significant acidosis/acidaemia
- Significant hyperkalaemia
- Significant hypokalaemia
- Significant hypermagnesaemia
- Significant hypomagnesaemia
- Significant hyperthermia
- Significant hypothermia
- Malignant hyperthermia
- Addisonian crisis
- Other (please specify)

#### 12. Most recent experience of perioperative cardiac arrest

* 19. What was the most likely **PRIMARY** cause of the perioperative cardiac arrest?

Choose single best option.

*If option not available, press 'previous' below for another category OR choose 'other' below.*

- Drug error
- Incompatible blood transfusion
- High neuraxial block
- Inadvertent neuraxial block during regional block
- Local anaesthetic toxicity (excessive dose and/or wrong route)
- Malignant hyperthermia
- Wrong gas supplied/unintentional connection to air
- Equipment failure
- Equipment lack
- Ventilator disconnection
- Other (please specify)

#### 13. Most recent experience of perioperative cardiac arrest

* 20. Did the patient survive the cardiac arrest?

*ROSC = return of spontaneous circulation*

- ◯ Died - no ROSC
- ◯ Died - transient ROSC (<20 min)
- ◯ Died - CPR stopped due to patient's known wishes (e.g. previous DNACPR decision or ReSPECT form)
- ◯ Survived cardiac arrest but died prior to hospital discharge
- ◯ Survived initial resuscitation and still in hospital
- ◯ Survived initial resuscitation but final outcome unknown
- ◯ Survived to hospital discharge
- ◯ Unknown

* 21. Were you present at the start of anaesthesia?

- ◯ Yes
- ◯ No
- ◯ Can't recall

* 22. Who was present in the room **at the start of anaesthesia**? Exclude yourself if you were present.

Tick all that apply.

- ☐ Consultant
- ☐ SAS doctor
- ☐ Post CCT or CESR doctor
- ☐ ST5+ or equivalent
- ☐ ST3-4 or equivalent
- ☐ CT2 or equivalent
- ☐ CT1 or equivalent – after Initial Assessment of Competence (IAC)
- ☐ CT1 or equivalent – before completion of Initial Assessment of Competence (IAC)
- ☐ Other (please specify)
- ☐ Anaesthesia Associate
- ☐ ODP/Anaesthetic nurse/Anaesthetic assistant
- ☐ Nurse/HCA e.g. scrub or recovery nurse
- ☐ Surgical team
- ☐ None
- ☐ Can’t recall
- ☐ Not applicable

* 23. List the number of anaesthesia providers (include yourself in the numbers) present **during the management** of the cardiac arrest?

| | 0 | 1 | 2 | 3 | ≥4 | Can't recall |
|---|---|---|---|---|---|---|
| Consultant | ○ | ○ | ○ | ○ | ○ | ○ |
| SAS doctor | ○ | ○ | ○ | ○ | ○ | ○ |
| Post CCT or CESR doctor | ○ | ○ | ○ | ○ | ○ | ○ |
| ST5+ or equivalent | ○ | ○ | ○ | ○ | ○ | ○ |
| ST3-4 or equivalent | ○ | ○ | ○ | ○ | ○ | ○ |
| CT2 or equivalent | ○ | ○ | ○ | ○ | ○ | ○ |
| CT1 or equivalent – after Initial Assessment of Competence (IAC) | ○ | ○ | ○ | ○ | ○ | ○ |
| CT1 or equivalent – before completion of Initial Assessment of Competence (IAC) | ○ | ○ | ○ | ○ | ○ | ○ |
| Anaesthesia Associate | ○ | ○ | ○ | ○ | ○ | ○ |
| ODP/Anaesthetic nurse/Anaesthetic assistant | ○ | ○ | ○ | ○ | ○ | ○ |

Other (please specify)

* 24. Who did you perceive to be leading the cardiac arrest?

* 25. Was a specific guideline used to assist in the management of perioperative cardiac arrest?

○ Yes

○ No

○ Can't recall

#### 14. Most recent experience of perioperative cardiac arrest

* 26. How was the specific guideline accessed? Tick all that apply.

☐ Smartphone

☐ Laminate

☐ In treatment pack

☐ Printed copy in theatre

☐ Computer/tablet

☐ Memory

☐ Can't recall

☐ Not applicable

☐ Other (please specify)

* 27. Was the theatre list or anaesthetic on-call shift stopped early?

- ◯ Yes – paused
- ◯ Yes – list stopped (includes cancelling remaining patients or transferring to care by a different team)
- ◯ No
- ◯ Not applicable (e.g. last case on list)
- ◯ Don't know
- ◯ Other (please specify)

* 28. Did any members of the team stand down from clinical activity* immediately after the event? Tick all that apply.

**does not include a break to document events or communicate with family, next of kin or other clinicians*

- ☐ Yes – I stood down
- ☐ Yes – some of the team
- ☐ Yes – all of the team
- ☐ No – because this was the end of the list or shift anyway
- ☐ No one stood down (e.g. continued with the next case)
- ☐ Can't recall

#### 15. Most recent experience of perioperative cardiac arrest

* 29. How did you or the team members stand down from clinical activity* immediately after the event? Tick all those that best describe this.

**A break does not include a break to document events or communicate with family, next of kin or other clinicians.*

- ☐ Took a short break* (e.g. <1 hour)
- ☐ Took a sustained break* (e.g. >1 hour)
- ☐ Theatre list terminated early
- ☐ Anaesthetic on-call shift terminated early
- ☐ Can't recall
- ☐ Not applicable
- ☐ Other (please specify)

* 30. How satisfied or dissatisfied were you with the quality of the management of the arrest?

| Very satisfied | Satisfied | Neither satisfied or dissatisfied | Dissatisfied | Very dissatisfied |
| --- | --- | --- | --- | --- |
| ○ | ○ | ○ | ○ | ○ |

Please provide reason(s) for your answer.

* 31. Who was the main member of the resuscitating team to directly communicate with the patient’s relatives / next of kin following the event?

- ○ Me
- ○ Another member of the team
- ○ Not applicable (e.g. no next of kin immediately available)
- ○ Can't recall

#### 16. Most recent experience of perioperative cardiac arrest.

* 32. Excluding yourself, who was the main team member to directly communicate with the patient’s relatives/next of kin following the event? Tick all that apply.

- ☐ Consultant anaesthetist
- ☐ Trainee anaesthetist
- ☐ SAS anaesthetist
- ☐ Consultant surgeon
- ☐ Trainee surgeon
- ☐ SAS surgeon
- ☐ Consultant from ICU
- ☐ ICU Trainee
- ☐ SAS from ICU
- ☐ Anaesthesia associate
- ☐ Nursing staff
- ☐ Physician
- ☐ Don't know
- ☐ Not applicable
- ☐ Other (please specify)

* 33. Was there a debrief relating to the case?

- ◯ Yes – I attended
- ◯ Yes – unable to attend (work duties)
- ◯ Yes – unable to attend (on leave)
- ◯ Yes – I was not invited
- ◯ Yes – I decided not to attend
- ◯ No, but there will be
- ◯ No, none planned
- ◯ Don’t know
- ◯ Other (please specify)

#### 17. Most recent experience of perioperative cardiac arrest

* 34. When did the debrief occur?

- ◯ Immediately after the event (hot debrief)
- ◯ Delayed period after the event (cold debrief)
- ◯ Both
- ◯ Can't recall
- ◯ Not applicable
- ◯ Other (please specify)

* 35. What type of debrief was carried out? Tick all that apply.

- ☐ Informal
- ☐ Formal (i.e. with a trained facilitator)
- ☐ One-to-one
- ☐ Group
- ☐ Trauma risk management (TRiM)
- ☐ Critical Incident Stress Debriefing (CISD)
- ☐ Don't know
- ☐ Not applicable
- ☐ Other (please specify)

* 36. Do you agree or disagree with the following statement?

**I was satisfied with the debrief process following the event.**

| Strongly agree | Agree | Neither agree or disagree | Disagree | Strongly disagree | Not applicable |
|---|---|---|---|---|---|
| ◯ | ◯ | ◯ | ◯ | ◯ | ◯ |

Please provide reason(s) for your answer.

* 37. How was the case (or will the case be) reviewed? Tick all that apply.

- ☐ Mortality and morbidity meeting
- ☐ Audit/ QI / governance meeting
- ☐ Internal investigation meeting e.g. root cause analysis
- ☐ Structured judgement/ mortality review
- ☐ Non-anaesthetic departmental meeting
- ☐ Other (please specify)
- ☐ Multi-specialty meeting, grand round or similar
- ☐ Serious incident framework
- ☐ No review has been performed or planned
- ☐ Don't know
- ☐ Not applicable

* 38. Was there an inquest or equivalent (e.g. Procurator Fiscal)?

- ◯ Yes
- ◯ Pending
- ◯ No
- ◯ Don't know
- ◯ Prefer not to say
- ◯ Not applicable

* 39. Was the case followed by legal proceedings?

- ◯ No
- ◯ Too early to know
- ◯ Yes
- ◯ Don’t know
- ◯ Prefer not to say
- ◯ Not applicable

* 40. Do you agree or disagree with the following statement?

**I feel satisfied with the way in which the case was followed up and reviewed.**

| Strongly agree | Agree | Neither agree or disagree | Disagree | Strongly disagree |
|---|---|---|---|---|
| ○ | ○ | ○ | ○ | ○ |

Please provide reason(s) for your answer.

* 41. What type of support have you received for this most recent case of perioperative cardiac arrest?

| | Yes | No | Prefer not to say | Not needed |
|---|---|---|---|---|
| Informal support (e.g. from colleagues) | ○ | ○ | ○ | ○ |
| Formal support from dedicated experienced senior anaesthetist | ○ | ○ | ○ | ○ |
| Formal hospital wellbeing support | ○ | ○ | ○ | ○ |

Other (please specify)

* 42. Has the cardiac arrest episode impacted on your ability to deliver patient care?

○ Yes
○ No
○ Not sure
○ Prefer not to say

#### 18. Most recent experience of perioperative cardiac arrest

* 43. How did the cardiac arrest episode impact your ability to deliver care?

#### 19. Career experience of perioperative cardiac arrest

**The following questions are about your CAREER EXPERIENCE of perioperative cardiac arrest.**

* 44. In your career, have any of your experiences of perioperative cardiac arrest (as the primary anaesthetist and those attended as a helper) had an **ADVERSE** impact on your **professional/work** life?

- ◯ Yes
- ◯ No
- ◯ Prefer not to say
- ◯ Not sure
- ◯ Not applicable - no prior experience of perioperative cardiac arrest

#### 20. Career experience of perioperative cardiac arrest

* 45. How did your experiences have an **adverse** impact on your **professional/work** life? Please tick all that apply.

- ☐ Time off work
- ☐ Loss of professional confidence
- ☐ Work related anxiety and stress
- ☐ Impacted relationship with colleagues
- ☐ Change in job plan
- ☐ Change hospital
- ☐ Change career
- ☐ Complaint about your performance
- ☐ Internal investigation about your performance
- ☐ Suspension
- ☐ GMC referral
- ☐ GMC investigation
- ☐ Civil Litigation
- ☐ Criminal investigation
- ☐ Prefer not to say
- ☐ Not applicable - no prior experience of perioperative cardiac arrest
- ☐ Other (please specify).

* 46. In your career, have any of your experiences of perioperative cardiac arrest (as the primary anaesthetist and those attended as a helper) had a **POSITIVE** impact on your **professional/work life**?

*If yes, please provide details below.*

- ◯ Yes
- ◯ No
- ◯ Not sure
- ◯ Prefer not to say
- ◯ Not applicable - no prior experience of perioperative cardiac arrest

If yes, please provide details

* 47. In your career, have any of your experiences of perioperative cardiac arrest (as the primary anaesthetist and those attended as a helper) had an **ADVERSE** impact on your **personal** life?

- Yes
- No
- Not sure
- Prefer not to say
- Not applicable - no prior experience of perioperative cardiac arrest

#### 21. Career experience of perioperative cardiac arrest

* 48. How did your experiences have an **adverse** impact on your **personal** life? Please tick all that apply.

- Impacted relationship with partner and/or children
- Impacted relationship with relatives
- Impacted relationship with friends
- Needed medical advice or care
- Other (please specify)
- Needed psychological support
- Prefer not to say
- Not applicable - no prior experience of perioperative cardiac arrest

* 49. In your career, have any of your experiences of perioperative cardiac arrest (as the primary anaesthetist and those attended as a helper) had a **POSITIVE** impact on your **personal life**?

*If yes, please provide details below.*

- Yes
- No
- Not sure
- Prefer not to say
- Not applicable - no prior experience of perioperative cardiac arrest

If yes, please provide details

#### 22. Demographics and workplace characteristics

**The following questions are about your current work practices.**

* 50. What gender do you identify yourself as?

- Female
- Male
- Other
- Prefer not to say

* 51. What is your age?

- <25 years old
- 25 – 35 years old
- 36 – 65 years old
- >65 years old
- Prefer not to say

* 52. What is your current role in Anaesthesia?

* 53. How long have you been an anaesthetist/anaesthesia associate? Please specify the number of years or months.

Years

Months

* 54. Do you currently work out of hours (weekend and/or nights)?

- Yes
- No

* 55. Do you have a subspecialty? Tick all that apply.

- [ ] Airway/ Head & neck
- [ ] Paediatrics
- [ ] Cardiothoracics
- [ ] Intensive care
- [ ] Neurosurgery
- [ ] Obstetrics
- [ ] Day case anaesthesia
- [ ] Orthopaedics
- [ ] Other (please specify)
- [ ] Bariatrics
- [ ] Transplant
- [ ] Trauma
- [ ] Regional anaesthesia
- [ ] Vascular
- [ ] Eyes
- [ ] None of the above
- [ ] Not applicable

* 56. What type of organisations do you currently work in?

- ( ) NHS
- ( ) Independent sector (non-NHS)
- ( ) Both

57. What country or region are you reporting from?

#### 23. Thank you for your contribution to the NAP7 baseline survey.

**Please contact your Local Coordinator that you have completed this survey.**
**You will then receive a certificate for completing the NAP7 Baseline Survey.**

**NAP7 starts on Wednesday 16th June 2021.**

**Please inform your Local Coordinator of all cases which may fulfil the criteria of perioperative cardiac arrest.**

**For more details please visit https://www.nationalauditprojects.org.uk/NAP7-Home#pt**
