## Supplementary material for "Comparing human vs. machine-assisted analysis to develop a new approach for Big Qualitative Data Analysis": Collaborator list

**Appendix S1 – Collaborator list**

S. Agarwal, Consultant, Department of Anaesthesia, Manchester University Hospitals Foundation Trust, Manchester, UK

D.C. Bouch, Consultant, Department of Critical Care and Anaesthesia, University Hospitals of Leicester NHS Trust, Leicester, UK

J. Cordingley, Consultant, Department of Critical Care and Anaesthesia, Barts Health NHS Trust, London, UK

L. Cortes, Royal College of Anaesthetists, London, UK

M. T. Davies, Consultant, Department of Critical Care and Anaesthesia, North West Anglia NHS Trust, UK

J. Dorey, Royal College of Anaesthetists, London, UK

Simon J. Finney, Consultant, Department of Critical Care and Anaesthesia, Barts Health NHS Trust

Gudrun Kunst Consultant and Professor of Anaesthesia, Department of Anaesthesia, Kings College Hospital London, School of Cardiovascular and Metabolic Medicine & Sciences, King’s College London

S.W. Kendall, Medical Director, North East and Yorkshire, NHS England and NHS Improvement, London, UK and Royal College of Surgeons representative

J. Lourtie, Royal College of Anaesthetists, London, UK

D. N. Lucas, Consultant, Department of Anaesthesia, London North West University Healthcare NHS Trust, London, UK

I. K. Moppet, Professor of Anaesthesia and Peri-operative Medicine, University of Nottingham, UK

R. Mouton, Consultant, Department of Anaesthesia and Intensive Care Medicine, Southmead Hospital, Bristol, UK

G. Nickols, Consultant, Department of Anaesthesia and Intensive Care Medicine, Southmead Hospital, Bristol, UK

J.P. Nolan, Professor, Resuscitation Medicine, Warwick Clinical Trials Unit, University of Warwick, Consultant, Department of Anaesthesia and Intensive Care Medicine, Royal United Hospitals Bath NHS Foundation Trust

Fiona C. Oglesby, Specialty Registrar, Bristol School of Anaesthesia, Severn Deanery, Bristol, UK

V. J. Pappachan, Consultant, Department of Paediatric Anaesthesia and Intensive Care Medicine, University Hospital Southampton NHS Foundation Trust, Southampton, UK

B. Patel, Royal College of Anaesthetists, London, UK

F. Plaat, Consultant, Department of Anaesthesia, Imperial College Healthcare NHS Trust, London, UK

K. Samuel, Consultant, Department of Anaesthesia and Intensive Care Medicine, Southmead Hospital, Bristol, UK

B. R. Scholefield, NIHR Clinician Scientist, Institute of Inflammation and Ageing, University of Birmingham, Birmingham, UK

J. H. Smith, Consultant, Department of Anaesthesia, Great Ormond Street Hospital, London, UK

C. Taylor, Royal College of Anaesthetists, London, UK

L. Varney, Anaesthesia Associate, Department of Anaesthesia, University College London Hospitals, London, UK

E.C. Wain, Associate Specialist, Department of Anaesthesia, Robert Jones and Agnes Hunt Orthopaedic Hospital, Gobowen, UK
